# Multivariate neural signatures of worry become more differentiated across adolescence and young adulthood: An ultra-high field (7T) fMRI study

**DOI:** 10.64898/2026.09.26.26364082

**Authors:** Cecilia A. Westbrook, Helmet T. Karim, Carmen Andreescu, Cecile D. Ladouceur

## Abstract

Worry is an important contributor to anxiety and depressive disorders, particularly in adolescence. Underlying changes in brain networks are thought to drive development of pathological worry, yet the degree to which neural correlates of worry vary by age is unknown. Here, we use multivariate pattern analysis (MVPA) to investigate individual variance in brain activity during worry in a cross-sectional sample of adolescents and adults varying in worry symptoms.

Participants (n=69; ages 12-31) completed a personalized worry-induction fMRI task. Brain activity was averaged across k=268 brain regions from the Shen atlas. Participants also completed self-reports assessing trait worry (PSWQ), anxiety (SCARED), and depression (MFQ) severity. A random-forest model was trained on estimated parameters from worry vs. neutral blocks using leave-one-subject-out cross-validation. Age and symptomatology measures were correlated with model performance metrics.

The overall cross-validated model had accuracy=66.71%, balanced accuracy=62.61%, sensitivity=80.43%, and specificity=44.78%. Trial-level accuracy increased with in-scanner worry ratings regardless of condition (*β* =0.62, *p*<.001). Accuracy (*r*=0.30, *p*=.01) and balanced accuracy (*r*=0.31, *p*=.01) significantly correlated with age but did not relate to symptom severity.

Brain patterns associated with worry become more differentiated across adolescence, suggesting that neural processes of worry consolidate across this period. Thus, early adolescence may be an opportune time for interventions to alter worry, but interventions adapted from adult populations may require tailoring for adolescent patients.

---

Anxiety disorders are the most common psychiatric disorders, with some of the greatest impacts on overall morbidity (GBD 2019 Mental Disorders Collaborators, 2022). This is especially true among adolescents, where the incidence of anxiety symptomatology has dramatically increased over the past two decades (Cybulski et al., 2021). Anxiety disorders— among many forms of psychopathology—most commonly onset in adolescence (Solmi et al., 2022) and place adolescents at increased risk for disrupted psychosocial function in adulthood (Essau et al., 2014) and greater risks for psychopathology including suicidality (Ferdinand and Verhulst, 1995; Kanwar et al., 2013). Yet adolescence is also a time of heightened psychological and neural plasticity (Luna et al., 2015), representing a window of opportunity for interventions to prevent or decrease anxiety symptoms.

A major contributor to anxiety symptomatology is worry, a complex cognitive-affective process comprising the tendency to engage in perseverative negative cognitions focused on future threats, and experience associated feelings of apprehension (Kircanski et al., 2015). Worry is a normative trait across the population, and at low levels is thought to facilitate problem-solving (Sweeny and Dooley, 2017); however, chronic and severe worry plays a causal role in development of anxiety disorders (Ehring and Behar, 2020), particularly generalized anxiety disorder (GAD) where it is a hallmark symptom (American Psychiatric Association, 2013). During adolescence, worry contributes to development of psychopathology including anxiety disorders and depression (Anniko et al., 2019; Tseliou et al., 2024), especially in girls, who develop anxiety disorders at around twice the rate of boys, with symptoms rising significantly during early adolescence (Beesdo-Baum and Knappe, 2012). However, the developmental mechanisms of perseverative cognition in this period are poorly understood.

Worry is measurable in children as young as 3 (Muris et al., 2002), but increases across childhood (Caes et al., 2016), developing in tandem with executive functions such as working memory and attentional control (Songco et al., 2020). During adolescence, the form and character of worry is thought to shift from concrete fears of childhood into more abstract and conjectural forms of thought (Vasey and Daleiden, 1994), mirroring the development of capacities for speculative thought about the future (Pine et al., 2021). Worry may also shift from mental imagery to a predominantly-verbal mode; although this was previously proposed to occur in middle childhood (Vasey, 1993), a recent review of evidence concluded that adolescence may instead be the period where worry shifts more into a “thinking-like process” (Songco et al., 2020).

Adolescence is a key developmental period for the neural processes driving these changes. Puberty initiates a cascade of reorganization and refinement of brain networks involved in executive functions (Larsen and Luna, 2018) and emotion regulation (Ladouceur, 2012). Subcortical reward- and emotion-related structures such as striatum and amygdala become more reactive compared to a relative dampening of regulatory activity in prefrontal cortex (PFC) (Houdé and Borst, 2022). Functional specialization in PFC and other brain networks increases, supporting the capacity for abstract thinking (Dumontheil, 2014). Together, these changes are likely reflected in gradual changes in the form and contents of worry, from more concrete, process-driven anxieties of childhood, to the abstract, verbal perseverative thoughts of adults (Pine et al., 2021). If so, this highlights the potential for interventions in adolescence to prevent worry *consolidation*. Furthermore, it implies that worry itself may look different in adolescents, which has implications for intervention as current theory is largely based on downward extensions of models developed for adult worry (Songco et al., 2020).

We previously examined neurodevelopmental correlates of worry using a worry-reappraisal paradigm which we have used in older-adult samples (Andreescu et al., 2016; Karim et al., 2025, 2017) in 69 individuals ranging in age from 12-31 using ultra high-field (7T) neuroimaging to focus on regions of interest within fronto-amygdala circuitry (Westbrook et al., 2026). We found that worry-induced activity in the amygdala—a region known to be altered in anxiety disorder psychopathology (Fox and Shackman, 2019)—was greater as a function of age. This finding supports the supposition that worry processes differ between adolescence and adulthood. Specifically, it raises the possibility that adolescents may use alternative or complementary networks to worry. Investigating this possibility, however, lies outside the abilities of conventional (univariate) neuroimaging analyses, which rely primarily on quantifying the magnitude of activation within specific regions or networks of interest (Davis et al., 2017). Although this approach provides valuable information about functions within specific brain areas, it does not capture patterns of activity among regions.

In order to examine the *similarity* of brain patterns across adolescence, we therefore turned to multivariate pattern analysis (MVPA), which leverages variance across the whole brain to identify distributed patterns predictive of specific outcomes or measures (Davis et al., 2014). This approach can out-perform univariate models; for instance, Keulers et al. (2012) found maturational effects across adolescents in multivariate brain activity during a go/no-go task despite finding very few small univariate results. Multivariate results can also show greater test-retest reliability than univariate results, meaning they are more robust to common concerns about replicability and over-fitting in fMRI (Kragel et al., 2021). MVPA approaches have demonstrated utility for diagnosis, prognosis and treatment prediction across a number of psychiatric disorders (Dwyer et al., 2018). Our MVPA model additionally benefits from the enhanced signal-to-noise ratio in this study, which is the first to utilize 7T to investigate the development of worry in adolescence.

Our overall analytic strategy was as follows: first, we would use MVPA to train a random-forest model on the previously-mentioned sample to attempt to discriminate worry vs. neutral trials. We would then extract the classification (i.e., discrimination) accuracy of those models and examine their relationship to age and other demographic variables. We hypothesized that 1) the model would predict worry vs. neutral trials with accuracy greater than chance, indicating that worry is a distinct mode of thought discriminable from neutral; 2) the model would be more likely to predict a trial is in the worry condition when participants self-reported higher levels of worry during that trial, indicating that participants are able to accurately self-report on their degree of worry; and 3) model accuracy and balanced accuracy would be greater with age, suggesting that adolescents use more variable brain networks than adults to worry. We did not specifically hypothesize relationships of model accuracy or balanced accuracy to sex but investigated this relationship as an exploratory analysis due to the known sex differences in anxiety. Finally, in addition to state worry induced during the task, we also conducted exploratory analyses of the relationship to trait measures of worry, anxiety and depression to investigate relationships to symptomatology in the real world.

## Method

We have previously published (Westbrook et al., 2026) a manuscript examining univariate analyses of the same dataset. Portions of the method and results including the task description and figure and demographics table are reproduced from that manuscript.

### Ethics Statement

All participants provided written informed consent or assent with parent or guardian for adolescents prior to participating in the study. Study procedures were approved by the University of Pittsburgh Institutional Review Board.

### Participants and Procedure

Participants (n=97) were adolescents (ages 12-17; 19 female) and young adults (ages 18-31; 31 female) from the greater Pittsburgh area, recruited on the basis of self-reported worry severity, with a goal of two-thirds of the sample reporting moderate-to-severe levels of worry (measured using the Penn State Worry Questionnaire for adults and children; see Methods).

Exclusion criteria included: presence of a neurological or serious medical condition, lifetime presence of psychotic or autistic spectrum disorder, MRI contraindications, medication that affects the central nervous system, pregnancy and breast-feeding.

Eligible participants were invited to complete a virtual visit, which comprised informed consent procedures and a brief interview to elicit personalized worry, and a single MRI study visit.

From 97 recruited participants, 69 (*N*=25 adolescents and *N*=44 adults) had complete data. Reasons for exclusion were: did not attend interview (*N*=2), did not attend scan (*N*=17), poor scanner coverage (*N*=2), excessive motion (*N*=1), technical issues during scan (*N*=3), missing questionnaire or demographic data (*N*=3). The MVPA models were trained including the 3 participants missing questionnaire/demographic data (*N*=72) because they had complete fMRI data but were excluded from final analyses.

### Measures

#### Assessments

The Penn State Worry Questionnaire (PSWQ; Meyer et al., 1990) and the PSWQ for children and adolescents (Chorpita et al., 1997) were used to assess levels of worry in adults and adolescents, respectively. The Screen for Child Anxiety Related Emotional Disorders (SCARED) for children and adolescents (Birmaher et al., 1997) and adults (Angulo et al., 2017) were used to assess anxiety symptoms and the Mood and Feelings Questionnaire (MFQ; Angold et al., 1995) was used to measure depression in both adults and adolescents. Participants (adult) or parents (adolescent) completed an in-house socio-demographic questionnaire which also included a question about sex assigned at birth. Adolescent PSWQ and SCARED self-reports were used for present analyses. PSWQ and SCARED scores were z-scored within age group (adolescent vs. adult) prior to combining for use in statistical tests.

#### fMRI Task and Instructions

The Worry Induction and Reappraisal Task is an fMRI task designed to elicit in-scanner personalized worry induction and worry reappraisal (Andreescu et al., 2016; Karim et al., 2025, 2017). The task contains three types of trials: Neutral (10), Worry induction (16), and worry Reappraisal (8). Neutral trials involve statements selected from a pool of non-worrisome, fact-based sentences (e.g., “There are 26 letters in the alphabet”). Worry and Reappraisal trials feature personalized statements. Prior to scanning, participants identified at least 16 personal worry induction statements via a semi-standardized interview. These were described as “a genuine worry or concern that you face on a day-to-day basis.” Participants then rated how worried they felt when reading each statement they identified, using a Likert scale from 1 to 5 (not worried at all – extremely worried). Next, participants worked with their interviewer to develop at least 8 worry reappraisal statements. Reappraisal was described as to “change the way you see or think about a situation so that it makes you feel better, less anxious.” For example, a worry induction statement could be “I worry that I will not get into my top choice for college and the rest of my career will fall apart,” and a worry reappraisal statement could be “My career will not fall apart if I do not get into my top choice for college, and I could still get into a really good college.”

During two ten-minute fMRI blocks, participants saw worry or neutral statements for 25 seconds. They were previously instructed to “read and think about” these statements. After 10 seconds, participants rated the level of worry from 1 (not worried at all) to 5 (extremely worried) using a five-finger MR-compatible glove. They had 15 seconds to rate/re-rate as many times as they wanted, followed by a 10-second fixation cross before the next statement appeared. Half of the worry statements were followed by a reappraisal statement selected to match it, during which participants had been instructed to decrease worry severity using reappraisal. This again was displayed for 25 seconds and rated at 10 seconds. Block order was semi-randomized to four scan orders to which participants were randomly assigned.

See Figure 1 for a visual depiction of the task.

**Figure 1.**
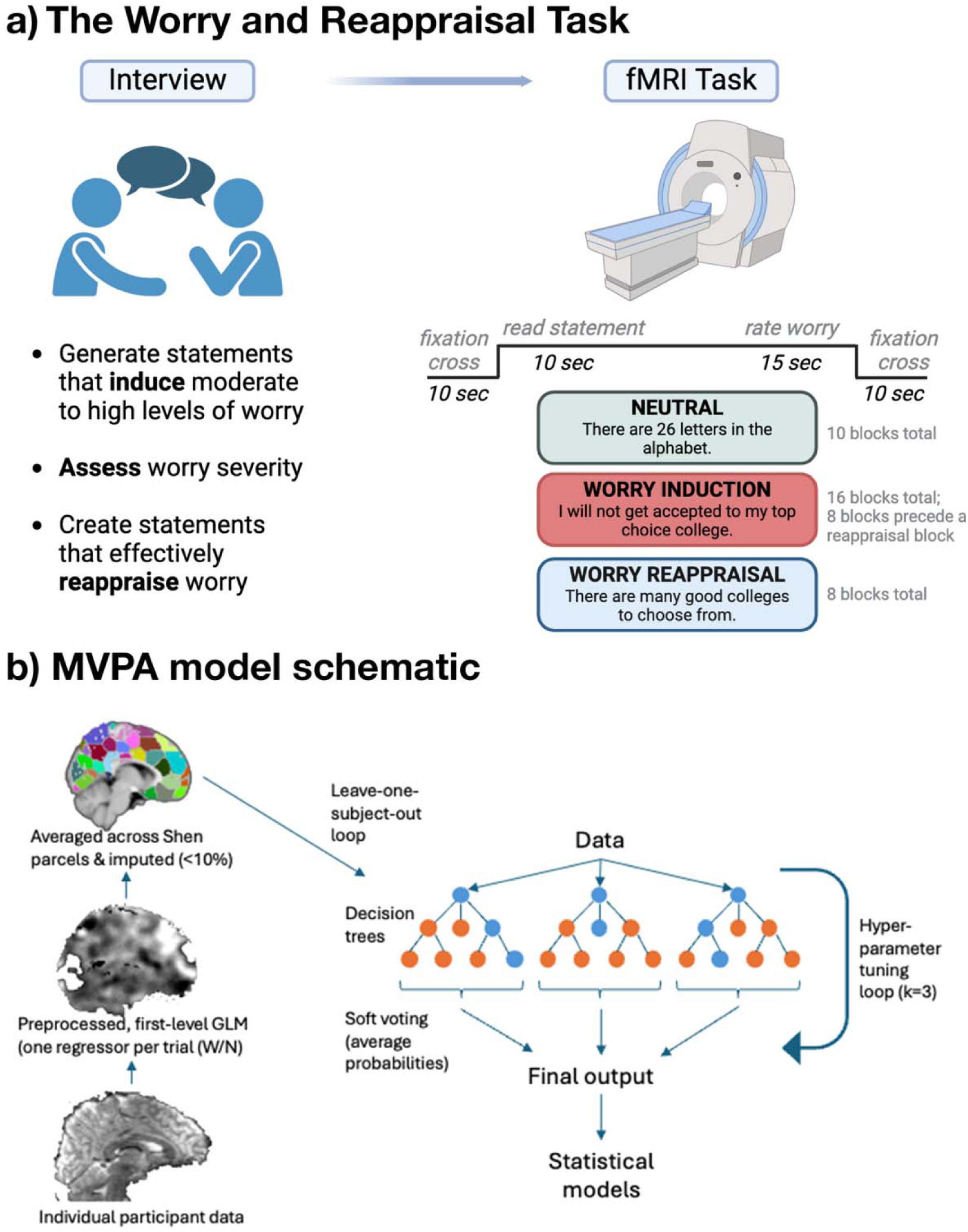
The Worry Induction and Reappraisal Task and MVPA model schematic. Panel a) depicts the Worry Induction and Reappraisal Task used in the study. Participants completed an interview with a trained study staff member to identify worry topics, reappraisal statements for these worry topics, and neutral statements. These were then used as stimuli in the task, during which participants were asked to worry as normal (Worry), reappraise that worry using their pre-generated reappraisal statements (Reappraisal), or think about a neutral topic as normal (Neutral). During the task they provided continuous ratings of their worry level on a 1-5 Likert scale. Panel b) depicts the overall model schematic for MVPA models. Individual participant data was preprocessed and analyzed with a level-1 univariate general linear model (GLM) to produce individual beta maps for each trial of each trial type of interest (e.g., Worry and Neutral) for each participant. These were then averaged across parcels from the Shen et al. (2013) atlas. Empty parcels arising from signal dropout (mean 2.3 parcels per participant, max 17/268, 0.9% of total data) were imputed using multiple regression via the imputer function from scikit-learn. A balanced random forest classifier was then trained on the 69 (subject) x 268 (parcel) dataset using leave-one-subject-out cross-validation.

### fMRI Data Collection and Analysis

#### Data collection and preprocessing

MR images were acquired on a 7T Siemens Magnetom scanner on a custom 16-transmit and 32-receive channels (Tic-Tac-Toe design) radiofrequency (RF) coil system (Ibrahim et al., 2013). Participants completed a practice session in a mock scanner prior to scanning. Structural images were acquired using a 3D multi-echo Magnetization Prepared Rapid Gradient Echo (MPRAGE) sequence (TR = 3000 ms, TE = 2.17 ms, TI = 1200 ms, flip angle = 8°, in-plane resolution of 230 ×320, 256 axial slices, 0.75 mm^3^ isotropic resolution, and GRAPPA acceleration factor of 2). Functional images were acquired using a T2*-weighted BOLD echoplanar imaging sequence with the following parameters: TR = 1000 ms, TE = 20 ms, flip angle = 65°, in-plane resolution of 110 × 110, 60 axial slices, 2 mm^3^ isotropic resolution, and multiband acceleration factor of 3.

Image preprocessing and data analyses were conducted primarily in SPM12 (Penny et al., 2011). MPRAGE were registered to MNI space and segmented into 6 tissue classes (Ashburner and Friston, 2005). Skull-stripping was completed by thresholding grey matter, white matter, and cerebrospinal fluid tissue probability maps by 0.1 and then conducting image filling and closing. Functional images were motion-corrected and skull-stripped using FSL’s BET. Skull-stripped functional images were registered to skull-stripped structural images, normalized into MNI space and smoothed at a FWHM kernel of 8mm. A 1/128-Hz bandpass filter was applied to account for scanner drift. Individual runs were excluded if >20% of volumes had head jerks as defined by the artifact repair toolbox (https://github.com/Remi-Gau/art_repair).

Regressors were constructed by convolving boxcar functions of the Worry, Reappraisal and Neutral conditions with the canonical hemodynamic response function. A single regressor was constructed for each trial in each condition in each run, resulting in 16 Worry regressors, 8 Reappraisal regressors, and 10 Neutral regressors. Nuisance regressors were constructed for 6 rigid-body motion parameters. Data were modeled using an autoregressive AR(1) model to account for biorhythms or other unmodeled activity. Contrasts were constructed for each regressor against unmodeled baseline, and the resulting beta maps were carried forward into MVPA analyses.

#### MVPA analyses

Analyses were conducted using scikit-learn version 1.5.0 (Kramer, 2016) and imbalanced-learn 0.12.4 (Lemaître et al., 2017) implemented in Python 3.10.12 (Python Software Foundation, Wilmington, Delaware), via jupyterlab version 4.2.4 (https://github.com/jupyterlab/jupyterlab). Individual beta maps for each condition were averaged across each of the 268 parcels from Shen et al. (2013) parcels arising from signal dropout (mean 2.3 parcels per participant, max 17/268, 0.9% of total data) were imputed using multiple regression. A balanced random forest classifier was trained on these brain maps to discriminate Worry vs. Neutral trials. A balanced random forest classifier is a random forest classifier which draws a bootstrap sample from the smaller class (Neutral) and samples the same number of items from the larger class (Worry) with replacement. Trials were classified across all subjects using leave-one-subject-out (LOSO) cross-validation for a total of 69 folds. Hyperparameters were fixed at 500 estimators and 16 features per split based on an initial randomized search (1,000 candidates per fold) which found near-uniform selection of parameters across folds. Two models were run: one which trained and tested on Worry and Neutral trials only, and one which trained on Worry and Neutral trials and tested on Worry, Neutral and Reappraisal trials. This latter model was run to examine factors that could influence the model to classify Reappraisal trials as Worry or Neutral conditions. Follow-up models were run for the Worry/Neutral only model to help characterize the results (see Results).

#### Analytic plan

Model performance was assessed via total (raw) accuracy and balanced accuracy as output by imbalanced-learn. Balanced accuracy was selected to account for the imbalance in trial numbers (16 Worry/10 Neutral). Significance was tested against a null generated by shuffling condition labels across 1,000 permutations.

Group-level analyses examined relationships of model indices to continuous variables (e.g., age). Model parameters were average accuracy, balanced accuracy, and computed values for sensitivity (true positive / [true positive + false negative]) and specificity (true negative / [true negative + false positive]) using Worry as the reference condition. Welch two-sample t-tests with estimated degrees of freedom were conducted to examine differences in model indices between groups (e.g., ethnicity). Trial-level effects, such as those examining in-scanner (i.e., state) worry, were examined using linear and binomial generalized mixed-effects models with a random intercept per participant using the lme4 package (Bates et al., 2015); simple slopes were calculated using emmeans (Lenth, 2020). 4 trials (0.2%) had missing ratings, so were excluded from these analyses. Analyses were completed using R version 4.5.0 (R Core Team, 2021) implemented in RStudio version 2025.0.5.0 (Posit team (2025)).

## Results

### Participants

There were no significant differences between adolescent and adult participants in sex, race or ethnicity (full demographic and clinical characteristics in Table 1). There were significant differences in family income (*U* = 1001.5, *p* < .001) and parental level of education (*U* = 705.5, *p* < .05).

**Table 1.** Demographic and clinical characteristics: adolescents vs. adults. *Note.* Abbreviations: PSWQ = Penn State Worry Questionnaire, MFQ = Mood and Feelings Questionnaire. PSWQ and SCARED scores were z-scored within age group (adolescent vs. adult) prior to combining for use in statistical tests.

|  | Adolescent |  | Adult |  |  |
| --- | --- | --- | --- | --- | --- |
| | Mean \| SD | | | | $t(p)$ |
| Age (years) | 15.08 | 2.15 | 23.80 | 3.29 | -13.30 (<.001) |
| | No. \| % | | | | $\chi^2(p)$ |
| Sex assigned at birth |  |  |  |  | 0.05 (0.83) |
| Female | 19 | 76.00 | 31 | 70.45 |  |
| Male | 6 | 24.00 | 13 | 29.55 |  |
| Ethnicity |  |  |  |  | 0.64 (0.42) |
| Not Hispanic or Latino | 21 | 84.00 | 41 | 93.18 |  |
| Hispanic or Latino | 4 | 16.00 | 3 | 6.82 |  |
| Race |  |  |  |  | 7.11 (0.07) |
| Black | 3 | 12.00 | 3 | 6.82 |  |
| Asian/Pacific Islander | 0 | 0.00 | 9 | 20.45 |  |
| White | 21 | 84.00 | 28 | 63.64 |  |
| Biracial | 1 | 4.00 | 4 | 9.09 |  |
|  |  |  |  |  | *U (p)* |
| Parent Highest Level of Education |  |  |  |  | 705.5 (0.04) |
| High school degree | 2 | 8.00 | 13 | 29.55 |  |
| Community college degree | 2 | 8.00 | 0 | 0.00 |  |
| Bachelor's degree | 7 | 28.00 | 24 | 54.54 |  |
| Master's degree | 7 | 28.00 | 5 | 11.36 |  |
| Doctorate | 5 | 20.00 | 2 | 4.55 |  |
| Not reported | 2 | 8.00 | 0 | 0.00 |  |
| Total family income per year |  |  |  |  | 1001.5 (<.001) |
| \$0-30,000 | 2 | 8.00 | 28 | 63.64 | |
| \$30,000-70,000 | 4 | 16.00 | 14 | 31.82 | |
| \$70,000-110,000 | 2 | 8.00 | 0 | 0.00 | |
| \$110,000-170,000 | 10 | 40.00 | 1 | 2.27 | |
| >\$170,000 | 7 | 28.00 | 1 | 2.27 | |
|  | Mean | SD | Mean | SD | *t (p)* |
| PSWQ (z-scored) | -0.04 | 1.03 | 0.07 | 1.01 | -0.38 (0.71) |
| MFQ (z-scored) | -0.07 | 1.00 | 0.03 | 1.03 | -0.44 (0.67) |

### Worry Ratings

The overall repeated-measures ANOVA comparing in-scanner worry ratings in Worry, Reappraisal and Neutral conditions was significant (*F(2,201)* = 20.08, *p* < .001). Post-hoc comparisons indicated that participants rated their worry higher during Worry (*M* = 3.40, *SD* = 0.78) than Neutral (*M* = 1.31, *SD* = 0.67; *t(68)* = 16.08, *p* < .001, *CI_diff_*= [1.82, 2.34]) and Reappraisal blocks (*M* = 2.17, *SD* = 0.62; *t(68)* = 11.52, *p* < .001, *CI_diff_* = [0.71, 1.01]), indicating that participants did worry during the worry blocks, and were able to successfully decrease their worry using reappraisal.

### Model performance across the full sample

The cross-validated model trained and tested on Worry and Neutral trials had accuracy = 66.71%, balanced accuracy = 62.61%, sensitivity = 80.43%, specificity = 44.78%. Both total and balanced accuracy exceeded all 1,000 label permutations (balanced accuracy: null *M* = 49.99%, *SD* = 0.87%, *p* < .001; total accuracy: null *M* = 57.29%, *SD* = *p* < .001).

### Model performance and demographic variables (Figure 2)

Participant age was significantly correlated with balanced accuracy (*r* = 0.31, *p* = .01) and total accuracy (*r* = 0.30, *p* < .05) (Figure 2a). There were no significant differences by sex or age by sex interactions. The relationship of balanced accuracy to age remained significant when adjusting for income (adjusted β = 0.32, *p* < .05), education (adjusted β = 0.31, *p* < .05), and race (adjusted β = 0.35, *p* < .01). In a linear mixed-effects model, there was a significant condition × age interaction on the model’s estimated probability that a trial was a Worry trial (β = 0.02, *p* = .001), such that the difference between Worry and Neutral trials increased with age (Figure 2b). In a binomial generalized mixed-effects model, the condition × age interaction on correct trial assignment was not significant (*p* = 0.38), indicating that discriminability improved with age for both Worry and Neutral trials rather than for one condition alone.

**Figure 2.**
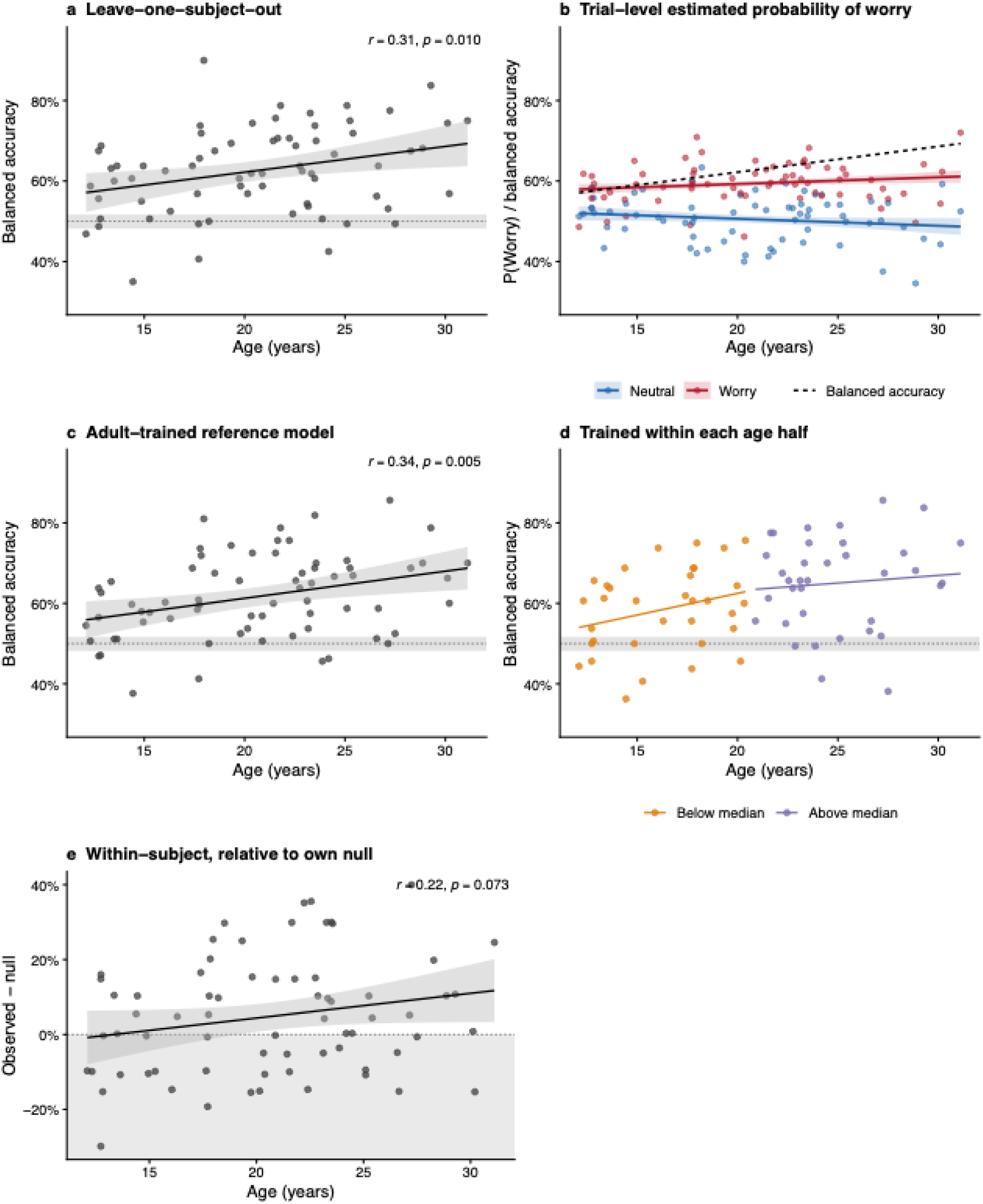
Model accuracy by age and comparison to follow-up models. Plots a and c-d depict per-participant balanced accuracy by age with a shaded band representing 95% confidence interval (CI). Horizontal shaded line and band represent the mean and 2.5-97.5^th^ percentile distribution of the permutation null; points falling above this line represent classification accuracy above chance. Panel a) depicts the primary leave-one-subject-out (LOSO) model; panel c) depicts the “frozen” reference version where this model was trained only on adult participants; panel d) depicts the age-stratified models, trained and tested separately on adolescents and adults; and panel d) depicts the within-subject model which was trained and tested within-participant using a leave-one-pair-out routine. Panel b) depicts the probability of the model classifying a trial as Worry or Neutral (P(Worry); model predictions from generalized linear model) as a function of age. Shaded bands represent 95% CI and dotted line overlay depicts the overall regression line for model accuracy by age.

#### Models stratified by age

To assess whether the model accuracy was biased towards adults, we split the sample at the median age (20.88 years) and retrained and tested the model within each group. The model in individuals below age 20.88 had accuracy = 63.64%, balanced accuracy = 58.30%, sensitivity = 81.34%, and specificity = 35.38%. The model in individuals above age 20.88 had accuracy = 68.22%, balanced accuracy = 64.93%, sensitivity = 79.25%, and specificity = 50.57% (Figure 2d).

#### “Frozen” reference model

To test whether model accuracy was due to younger adolescents having older training sets, we trained a new model on adult data only (*N* = 44), such that each adolescent’s training set was identical. Mean balanced accuracy for this model was 61.67%. The correlation with age remained significant for all participants (*r* = 0.34, *p* < .01) and the adolescent subset (*r* = 0.44, *p* < .05) (Figure 2c).

#### Within-subject model

To test whether adolescents had stable but idiosyncratic Worry representations, we trained a within-subjects model. For each of 10 iterations (one per Neutral trial; Worry trials selected without replacement), one pair of Worry and Neutral trials was held out and the remaining set was standardized and feature-reduced using principal components analysis, then subjected to training using logistic regression with three-fold cross-validation. Mean within-subject balanced accuracy was 54.78%, compared with a mean null of 49.97% generated by 1,000 permutations of condition labels (*z* = 3.60, *p* < .001). Within-subject accuracy was not associated with age (*r* = .19, *p* = .114) (Figure 2e).

#### Relationship of age to motion

To rule out a possible confound in the relationship with age and model accuracy, we examined the relationship of age to fourteen motion metrics across both runs (maximum translation, translation range, average scan-to-scan displacement, total translation, root-mean square motion, and number of head jerks at two thresholds). None of these relationships were significant (all *p* > 0.05). Coverage dropout requiring imputation was likewise unrelated to age (*r* = 0.17, *p* = 0.16).

### Model performance and clinical variables

There were no significant relationships between accuracy or balanced accuracy and SCARED, PSWQ or MFQ scores. Generalized linear mixed models examining classification by trial were similarly not significant. These were likewise not significant when analyzed separately in adolescents and adults, and when an interaction term for age was included.

### Model performance and worry rating

In a binomial generalized linear mixed model, the model was found to be significantly more likely to classify a trial as Worry as the worry rating of that trial increased (β = 0.62, *p* < .001). When an interaction term for the true condition (Worry vs. Neutral) was included, this was also significant (condition × rating *β* = -0.53, *p* < .05), indicating that the effect was driven by Neutral trials, such that Neutral trials where worry was rated higher were more likely to be classified as Worry trials. A parallel model on correct trial assignment showed the same pattern (rating β = -0.64, *p* < .001; condition × rating β = 0.63, *p* < .01). At the participant level, higher mean worry rating during Neutral blocks predicted lower overall classification performance (β = -.38, *p* < .01).

These results are visualized in Figure 3.

**Figure 3.**
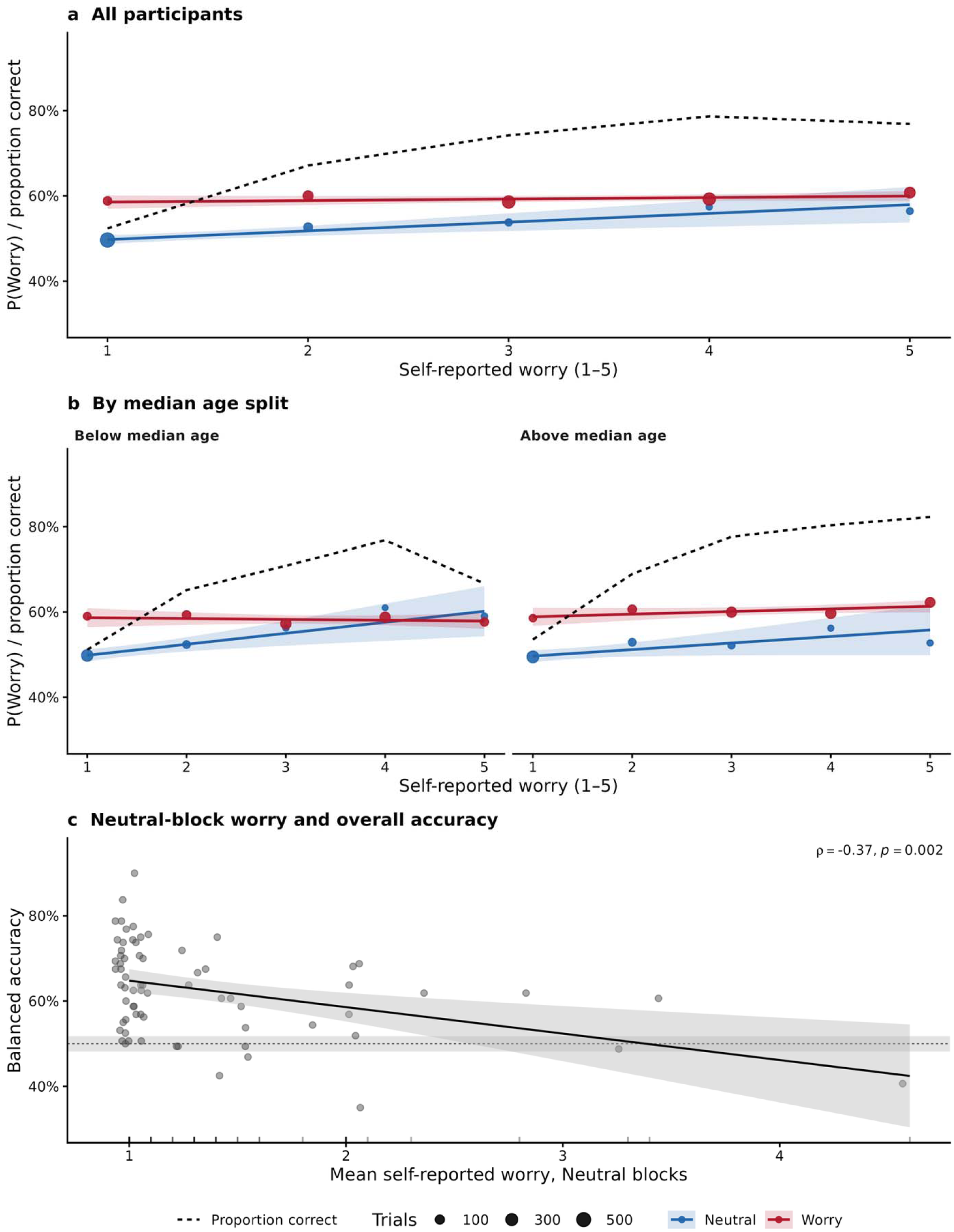
Model accuracy and probability of assigning worry by worry rating. Panel a) depicts the model-assigned probability of a trial being classified as Worry (P(Worry)) as a function of self-reported worry rating on that trial. Dashed line represents average proportion correct. Panel b) depicts proportion of correctly classified trials in the models trained only on the older and younger half of participants, classified as above-median (20.88 years) and below-median, respectively. Panel c) depicts the between-subjects balanced accuracy as a function of the mean worry each participant reported during Neutral trials. Regression line and 95% CI are plotted demonstrating that the model was less accurate for individuals who reported higher levels of worry during Neutral trials. Due to the skewed distribution, Spearman correlation was computed and was also significant (*ρ* = -0.37, *p* < .01) indicating that the relationship was not driven by influential outliers.

### Reappraisal classification and worry rating

In the model trained on Worry and Neutral and tested on Reappraisal trials, Reappraisal trials were classified as Worry on 75.1% of trials, compared with 80.4% of Worry (*M_diff_* = -5.3%, *p* < .05) and 55.2% of Neutral (*M_diff_* = 19.9%, *p* < .001) trials. In a binomial generalized linear mixed model, there was no significant relationship between worry rating and likelihood of classifying Reappraisal trials as Worry vs. Neutral (β = 0.12, *p* = 0.36).

### Feature importance by age

To examine differences in networks utilized for worry across age, we correlated feature importance for each of the 268 Shen features (calculated for each subject) with age. We divided the 268 parcels into networks by assigning each to one of the 7 networks identified by Yeo et al., 2011 (Yeo et al., 2011). The resulting correlations were near zero across networks, ranging from -0.04 to 0.03. Across the parcels with network assignments, none of the correlations survived Benjamini-Hochberg multiple comparisons correction.

## Discussion

Our primary finding in this cross-sectional MVPA investigation was that the accuracy of our random-forest model increased with age. This indicates that the patterns of brain activity corresponding to worry and neutral blocks were less differentiable in young adolescents and become more differentiable in adulthood. Two control models demonstrated that this finding did not simply arise from bias towards older participants: a stratified model trained only within adolescents performed worse than the model trained only in adults, and a model trained only on the adult sample reproduced the accuracy relationship with age across adolescence. And, although we cannot rule out unmeasured confounds, we did not find evidence for a correlation with age and head motion, which increases our confidence in the age-accuracy correlation as a true effect.

MVPA models can classify based on any features in the data, so we implemented follow-up measures to help characterize the discrimination performance in our sample. First, although overall classification was more accurate on trials with higher in-scanner worry ratings, the model was more likely to incorrectly assign label Neutral trials as Worry when they had higher worry ratings, and individuals who rated higher worry during Neutral trials had overall lower accuracy. Thus, the model appears to have captured *worry-related brain activity* specifically, and not simply task structure. However, the absence of a condition-specific age effect indicates that neither condition became more identifiable on its own; rather, the two became more distinct from one another across adolescence.

Examining the specific features that were most important for classification, we found that they were widely distributed across the brain. Thus, although adult patterns corresponding to worry were more consistent than those in adolescents, there was not one specific network that contributed more to worry. We note that classifier weights reflect noise suppression as well as signal, are unstable among correlated features, and are not directly comparable across models that differ in overall accuracy, and thus absence of relationships between worry and specific networks is not definitive evidence against anatomical specificity. However, this result does suggest that the correlation between age and model accuracy does not arise from adolescents recruiting more or less activity in specific regions, but rather from different suites of brain networks that coalesce into a more common pattern in adulthood. One interpretation of this trajectory is that these patterns are more variable, and adolescents gradually shift from idiosyncratic thought patterns, unique to each individual, to a more common worry pattern. If this were the case, however, we might have expected higher accuracy from our within-subjects models, but they performed only a little better than chance. Alternatively, it may indicate that the brain networks adolescents use to engage in worry and neutral thought have more overlap. This interpretation is somewhat supported by a trial-wise mixed model showing that classification improved equally at predicting Worry *and* Neutral trials across age.

We were surprised to not see feature importance concentrated in areas previously associated with worry, such as the default-mode network (DMN) (Makovac et al., 2020) or limbic brain regions such as the amygdala (e.g., Makovac et al., 2016; Wu et al., 2019).A highly-speculative explanation for these findings is that what differentiates adult from adolescent worry is the recruitment of specialized networks such as the DMN and amygdala-centric fear networks (e.g., Ellard et al., 2017; Feurer et al., 2021; Makovac et al., 2016), that such networks are less critical for the phenomenology of worry as captured in clinical measures, and perhaps serve other purposes such as recruiting autonomic systems to support physical symptoms of worry (Makovac et al., 2016). In support of this hypothesis, we showed in these same data that worry evoked recruitment of the paraventricular nucleus (PVN) of the hypothalamus, a region critical for HPA axis involvement (Westbrook et al., 2026), which we have also previously linked to worry in older adults (Andreescu et al., 2016, 2015; Kolobaric et al., 2022). In the absence of behavioral or cognitive correlates of this shift, this remains a speculative interpretation; future investigation using within-subjects longitudinal designs could perhaps illuminate the significance of this change. Nevertheless, it does suggest that other brain networks which we (Karim et al., 2025, 2017) have previously found during worry induction may warrant greater investigation.

Interestingly, we did not see relationships between model accuracy and clinical symptomatology. Although this may reflect our sample size being insufficient to detect questionnaire differences, it may also suggest that the increase in differentiability with age represents changes in the *process* of worry, but not in the distress or clinical impact (as reported via questionnaire) it causes. Another unexpected dissociation between neural activity and subjective experience came from the Reappraisal trials. A model trained on Worry and Neutral, which did not ever see Reappraisal trials, assigned about 75% percent of them to the Worry category. This is in spite of the fact that average worry rating dropped 1.23 points (*p* < .001) during Reappraisal trials. In fact, classification of Reappraisal trials was not related to worry rating during those trials at all. A potential explanation is that Reappraisal does not change or suppress the distributed representation of worry in the brain, but instead changes the subjective character of worry, that is, participants’ cognitive experience of their own thoughts, as we have previously argued about acceptance-based worry disengagement (Westbrook et al., 2023). In that case, the neural representation of reappraisal would be more similar to worry than to neutral thought. Or it may act as an add-on module, which could substantially increase cognitive load. Further research examining these possibilities would have significant clinical implications, as reappraisal is a widely-taught clinical skill and a core component of cognitive-behavioral therapy (Kazantzis et al., 2018).

Our study must be interpreted in light of its strengths and weaknesses, which we have described previously (Westbrook et al., 2026). There were several demographic differences between our adult and adolescent groups: most notably, there were fewer boys in the adolescent sample (although this difference did not reach statistical significance), and boys in this study reported lower levels of worry when viewing their worry topics in the scanner. This reduced worry may have impacted accuracy of the model for younger adolescent boys, although they represented a minority of the sample. This study also suffered from significant attrition due to being conducted during the early portion of the COVID-19 pandemic; we did not find significant demographic differences between those who did and did not complete the study.There were differences in income and education level between groups; we note that income was caregiver-reported for adolescents and self-reported for adults, which could partially account for the difference as young adults likely have lower incomes than full families. This study was not powered to thoroughly investigate socioeconomic differences; however, we note that our results were robust when controlled for income and parental education. Finally, this study utilized a cross-sectional design, which precludes studying within-subject neurodevelopmental changes with age. Nevertheless, this study had considerable strengths. We included participants across a wide age range when worry is known to undergo active development, and the sample was enriched such that 2/3 of participants reported moderate to severe worry in the past 2 weeks. Furthermore, it utilized ultra-high-field (7T) neuroimaging to enhance the spatial resolution, which is important for detecting neural activity in smaller subcortical regions.

Taken together, our results suggest that worry evolves across adolescence, with adolescents using different brain networks to worry, despite reporting similar levels of distress and resulting symptomatology. This has important clinical implications: on the one hand, it represents a window of opportunity, as we have evidence worry has not fully coalesced into its adult form in adolescence, and may thus be subject to intervention. On the other hand, it implies that interventions designed to treat and prevent worry in adults may target brain networks that are less relevant to adolescents. Future research aimed at identifying which networks are specific to adolescent worry, and how best to measure and target such brain networks, could provide fruitful avenues towards developing interventions specific to this age range.

## Data and code availability

Data for this study are publicly available on the NIMH Data Archive. Code for this project is publicly available at https://github.com/ceciwestbrook/AdolescentWorryMVPA

## Author contributions

Westbrook: Conceptualization, formal analysis, methodology, writing (original draft), writing (review & editing); Karim: Conceptualization, formal analysis, methodology, writing (review & editing); Andreescu: Funding acquisition, investigation, project administration, resources, writing (review & editing); Ladouceur: Conceptualization, funding acquisition, investigation, project administration, resources, supervision, writing (review & editing).

## Funding sources

This work was supported by the National Institute of Mental Health [grant numbers R21MH122808, R01MH108509]; and the Clinical and Translational Science Institute at the University of Pittsburgh [grant number UL1TR001857]. The funders had no role in study design, data collection, analysis or interpretation, writing of the report, or the decision to submit the article for publication.

## Declaration of competing interests

The authors have no conflicts of interest to disclose related to this project.

## Declaration of generative AI and AI-assisted technologies in the manuscript preparation process

During the preparation of this work the authors used ChatGPT (4.0 and 5.0) and Claude Opus 5 in order to assist with code production in Python and R. After using these tools, the authors reviewed and edited the content as needed and take full responsibility for the content of the published article.

## Notes

### Competing Interest Statement

The authors have declared no competing interest.

### Author Declarations

Study procedures were approved by the University of Pittsburgh Institutional Review Board.

## References

American Psychiatric Association, 2013. Diagnostic and statistic manual of mental disorders (5th ed., text rev.). Washington, D.C.:Publisher.

Andreescu, C., Mennin, D., Tudorascu, D., Sheu, L.K., Walker, S., Banihashemi, L., Aizenstein, H., 2015. The many faces of anxiety-neurobiological correlates of anxiety phenotypes. Psychiatry Research: Neuroimaging 234, 96–105. 10.1016/j.pscychresns.2015.08.013

Andreescu, C., Sheu, L.K., Tudorascu, D., Gross, J.J., Walker, S., Banihashemi, L., Aizenstein, H., 2016. Emotion reactivity and regulation in late-life generalized anxiety disorder: Functional connectivity at baseline and post-treatment. Am J Geriatr Psychiatry 24, 1040–1050. 10.1016/j.jagp.2016.07.015

Angold, A., Costello, E.J., Messer, S.C., Pickles, A., Winder, F., Silver, D., 1995. Development of a short questionnaire for use in epidemiological studies of depression in children and adolescents. International Journal of Methods in Psychiatric Research 5, 237–249.

Angulo, M., Rooks, B.T., Gill, M., 2017. Psychometrics of the screen for adult anxiety related disorders (SCAARED)-A new scale for the assessment of DSM-5 anxiety disorders. Psychiatry Research 253, 84–90. 10.1016/j.psychres.2017.02.034

Anniko, M.K., Boersma, K., Tillfors, M., 2019. Sources of stress and worry in the development of stress-related mental health problems: A longitudinal investigation from early-to mid-adolescence. Anxiety, Stress, & Coping 32, 155–167. 10.1080/10615806.2018.1549657

Ashburner, J., Friston, K.J., 2005. Unified segmentation. NeuroImage 26, 839–851. 10.1016/j.neuroimage.2005.02.018

Bates, D., Mächler, M., Bolker, B., Walker, S., 2015. Fitting Linear mixed-effects models using lme4. Journal of Statistical Software 67, 1–48. 10.18637/jss.v067.i01

Beesdo-Baum, K., Knappe, S., 2012. Developmental epidemiology of anxiety disorders. Child and Adolescent Psychiatric Clinics 21, 457–478. 10.1016/j.chc.2012.05.001

Birmaher, B., Khetarpal, S., Brent, D., Cully, M., Balach, L., Kaufman, J., Neer, S.M., 1997. The screen for child anxiety related emotional disorders (SCARED): Scale construction and psychometric characteristics. Journal of the American Academy of Child & Adolescent Psychiatry 36, 545–553. 10.1097/00004583-199704000-00018

Caes, L., Fisher, E., Clinch, J., Tobias, J.H., Eccleston, C., 2016. The development of worry throughout childhood: Avon Longitudinal Study of Parents and Children data. British Journal of Health Psychology 21, 389–406. 10.1111/bjhp.12174

Chorpita, B.F., Tracey, S.A., Brown, T.A., Collica, T.J., Barlow, D.H., 1997. Assessment of worry in children and adolescents: An adaptation of the Penn State Worry Questionnaire. Behaviour Research and Therapy 1997;35(6):569–581. 10.1016/S0005-7967(96)00116-7

Cybulski, L., Ashcroft, D.M., Carr, M.J., Garg, S., Chew-Graham, C.A., Kapur, N., Webb, R.T., 2021. Temporal trends in annual incidence rates for psychiatric disorders and self-harm among children and adolescents in the UK, 2003–2018. BMC Psychiatry 21, 229. 10.1186/s12888-021-03235-w

Davis, T., Goldwater, M., Giron, J., 2017. From concrete examples to abstract relations: The rostrolateral prefrontal cortex integrates novel examples into relational categories. Cerebral Cortex 27, 2652–2670. 10.1093/cercor/bhw099

Davis, T., LaRocque, K.F., Mumford, J.A., Norman, K.A., Wagner, A.D., Poldrack, R.A., 2014. What do differences between multi-voxel and univariate analysis mean? How subject-, voxel-, and trial-level variance impact fMRI analysis. NeuroImage 97, 271–283. 10.1016/j.neuroimage.2014.04.037

Dumontheil, I., 2014. Development of abstract thinking during childhood and adolescence: The role of rostrolateral prefrontal cortex. Developmental Cognitive Neuroscience 10, 57–76. 10.1016/j.dcn.2014.07.009

Dwyer, D.B., Falkai, P., Koutsouleris, N., 2018. Machine learning approaches for clinical psychology and psychiatry. Annual Review of Clinical Psychology 14, 91–118. 10.1146/annurev-clinpsy-032816-045037

Ehring, T., Behar, E., 2020. Transdiagnostic view on worrying and other negative mental content, in: Generalized Anxiety Disorder and Worrying. John Wiley & Sons, Ltd, pp. 43–68. 10.1002/9781119189909.ch4

Ellard, K.K., Barlow, D.H., Whitfield-Gabrieli, S., Gabrieli, J.D.E., Deckersbach, T., 2017. Neural correlates of emotion acceptance vs worry or suppression in generalized anxiety disorder. Social Cognitive and Affective Neuroscience 12, 1009–1021. 10.1093/scan/nsx025

Essau, C.A., Lewinsohn, P.M., Olaya, B., Seeley, J.R., 2014. Anxiety disorders in adolescents and psychosocial outcomes at age 30. Journal of Affective Disorders 163, 125–132. 10.1016/j.jad.2013.12.033

Ferdinand, R.F., Verhulst, F.C., 1995. Psychopathology from adolescence into young adulthood: An 8-year follow-up study. The American Journal of Psychiatry 152, 1586–94.

Feurer, C., Jimmy, J., Chang, F., Langenecker, S.A., Phan, K.L., Ajilore, O., Klumpp, H., 2021. Resting state functional connectivity correlates of rumination and worry in internalizing psychopathologies. Depression and Anxiety 38, 488–497. 10.1002/da.23142

Fox, A.S., Shackman, A.J., 2019. The central extended amygdala in fear and anxiety: Closing the gap between mechanistic and neuroimaging research. Neuroscience Letters, Functional Neuroimaging of the Emotional Brain 693, 58–67. 10.1016/j.neulet.2017.11.056

GBD 2019 Mental Disorders Collaborators, 2022. Global, regional, and national burden of 12 mental disorders in 204 countries and territories, 1990–2019: a systematic analysis for the Global Burden of Disease Study 2019. The Lancet Psychiatry 9, 137–150. 10.1016/S2215-0366(21)00395-3

Houdé, O., Borst, G., 2022. The Cambridge Handbook of Cognitive Development. Cambridge University Press.

Ibrahim, T., Zhao, Y., Krishnamurthy, N., Raval, S.B., Zhao, T., Wood, S., Kim, J., 2013. 20-To-8 channel Tx array with 32-channel adjustable receive-only insert for 7T head imaging. Presented at the International Society for Magnetic Resonance in Medicine, Salt Lake City, UT.

Kanwar, A., Malik, S., Prokop, L.J., Sim, L.A., Feldstein, D., Wang, Z., Murad, M.H., 2013. The association between anxiety disorders and suicidal behaviors: A systematic review and meta-analysis. Depression and Anxiety 30, 917–929. 10.1002/da.22074

Karim, H.T., Gross, J.J., Mennin, D., Gerlach, A., Tudorascu, D.L., Butters, M.A., Ladouceur, C., Andreescu, C., 2025. Older and wiser? The neural correlates of worry induction and reappraisal in older adults. Neuropsychopharmacol. 1–10. 10.1038/s41386-025-02193-1

Karim, H.T., Tudorascu, D.L., Butters, M.A., Walker, S., Aizenstein, H.J., Andreescu, C., 2017. In the grip of worry: cerebral blood flow changes during worry induction and reappraisal in late-life generalized anxiety disorder. Translational Psychiatry 7, e1204–e1204. 10.1038/tp.2017.180

Kazantzis, N., Luong, H.K., Usatoff, A.S., Impala, T., Yew, R.Y., Hofmann, S.G., 2018. The processes of cognitive behavioral therapy: A review of meta-analyses. Cogn Ther Res 42, 349–357. 10.1007/s10608-018-9920-y

Keulers, E.H.H., Goulas, A., Jolles, J., Stiers, P., 2012. Maturation of task-induced brain activation and long range functional connectivity in adolescence revealed by multivariate pattern classification. NeuroImage 60, 1250–1265. 10.1016/j.neuroimage.2011.12.079

Kircanski, K., Thompson, R.J., Sorenson, J.E., Sherdell, L., Gotlib, I.H., 2015. Rumination and worry in daily life: Examining the naturalistic validity of theoretical constructs. Clinical Psychological Science 3, 926–939. 10.1177/2167702614566603

Kolobaric, A., Karim, H.T., Banihashemi, L., Mizuno, A., Aizenstein, H.J., Andreescu, C., 2022. Are all anxieties created equal? Stress-related networks and anxiety phenotypes in old age. The American Journal of Geriatric Psychiatry 30, 801–812. 10.1016/j.jagp.2021.12.007

Kragel, P.A., Han, X., Kraynak, T.E., Gianaros, P.J., Wager, T.D., 2021. Functional MRI can be highly reliable, but it depends on what you measure: A commentary on Elliott et al. (2020). Psychol Sci 32, 622–626. 10.1177/0956797621989730

Kramer, O., 2016. Scikit-Learn, in: Kramer, O. (Ed.), Machine Learning for Evolution Strategies. Springer International Publishing, Cham, pp. 45–53. 10.1007/978-3-319-33383-0_5

Ladouceur, C.D., 2012. Neural systems supporting cognitive-affective interactions in adolescence: the role of puberty and implications for affective disorders. Front. Integr. Neurosci. 6. 10.3389/fnint.2012.00065

Larsen, B., Luna, B., 2018. Adolescence as a neurobiological critical period for the development of higher-order cognition. Neuroscience & Biobehavioral Reviews 94, 179–195. 10.1016/j.neubiorev.2018.09.005

Lemaître, G., Nogueira, F., Aridas, C.K., 2017. Imbalanced-learn: A python toolbox to tackle the curse of imbalanced datasets in machine learning. Journal of Machine Learning Research 18, 1–5.

Lenth, R., 2020. emmeans: Estimated Marginal Means, aka Least-Squares Means.

Luna, B., Marek, S., Larsen, B., Tervo-Clemmens, B., Chahal, R., 2015. An integrative model of the maturation of cognitive control. Annual Review of Neuroscience 38, 151–170. 10.1146/annurev-neuro-071714-034054

Makovac, E., Fagioli, S., Rae, C.L., Critchley, H.D., Ottaviani, C., 2020. Can’t get it off my brain: Meta-analysis of neuroimaging studies on perseverative cognition. Psychiatry Research: Neuroimaging 295, 111020. 10.1016/j.pscychresns.2019.111020

Makovac, E., Meeten, F., Watson, D.R., Herman, A., Garfinkel, S.N., D. Critchley, H., Ottaviani, C., 2016. Alterations in Amygdala-Prefrontal Functional Connectivity Account for Excessive Worry and Autonomic Dysregulation in Generalized Anxiety Disorder. Biological Psychiatry, Stress, Fear, and Anxiety 80, 786–795. 10.1016/j.biopsych.2015.10.013

Meyer, T.J., Miller, M.L., Metzger, R.L., Borkovec, T.D., 1990. Development and validation of the Penn State Worry Questionnaire. Behaviour Research and Therapy 28, 487–495. 10.1016/0005-7967(90)90135-6

Muris, P., Merckelbach, H., Ollendick, T., King, N., Bogie, N., 2002. Three traditional and three new childhood anxiety questionnaires: their reliability and validity in a normal adolescent sample. Behaviour Research and Therapy 40, 753–772. 10.1016/S0005-7967(01)00056-0

Penny, W.D., Friston, K.J., Ashburner, J.T., Kiebel, S.J., Nichols, T.E., 2011. Statistical parametric mapping: the analysis of functional brain images. Elsevier.

Pine, D.S., Wise, S.P., Murray, E.A., 2021. Evolution, emotion, and episodic engagement. AJP 178, 701–714. 10.1176/appi.ajp.2020.20081187

R Core Team, 2021. R: A language and environment for statistical computing. R Foundation for Statistical Computing, Vienna, Austria.

Shen, X., Tokoglu, F., Papademetris, X., Constable, R.T., 2013. Groupwise whole-brain parcellation from resting-state fMRI data for network node identification. NeuroImage 82, 403–415. 10.1016/j.neuroimage.2013.05.081

Solmi, M., Radua, J., Olivola, M., Croce, E., Soardo, L., Salazar de Pablo, G., Il Shin, J., Kirkbride, J.B., Jones, P., Kim, J.H., Kim, J.Y., Carvalho, A.F., Seeman, M.V., Correll, C.U., Fusar-Poli, P., 2022. Age at onset of mental disorders worldwide: large-scale meta-analysis of 192 epidemiological studies. Mol Psychiatry 27, 281–295. 10.1038/s41380-021-01161-7

Songco, A., Hudson, J.L., Fox, E., 2020. A cognitive model of pathological worry in children and adolescents: A systematic review. Clin Child Fam Psychol Rev 23, 229–249. 10.1007/s10567-020-00311-7

Sweeny, K., Dooley, M.D., 2017. The surprising upsides of worry. Social and Personality Psychology Compass 11.

Tseliou, F., Riglin, L., Thapar, A.K., Heron, J., Dennison, C.A., Armitage, J.M., Thapar, A., Rice, F., Collishaw, S., 2024. Childhood correlates and young adult outcomes of trajectories of emotional problems from childhood to adolescence. Psychological Medicine 54, 2504–2514. 10.1017/S0033291724000631

Vasey, M.W., 1993. Development and cognition in childhood anxiety: The example of worry. Advances in Clinical Child Psychology 15, 1–39.

Vasey, M.W., Daleiden, E.L., 1994. Worry in children, in: Worrying: Perspectives on Theory, Assessment and Treatment, Wiley Series in Clinical Psychology. John Wiley & Sons, Oxford, England, pp. 185–207.

Westbrook, C., Kolobaric, A., Karim, H.T., Pine, D.S., Andreescu, C., Ladouceur, C.D., 2026. Identifying neurodevelopmental signatures of worry: An ultra-high field (7-Tesla) fMRI study among adolescents and young adults. Developmental Cognitive Neuroscience 79, 101718. 10.1016/j.dcn.2026.101718

Westbrook, C.A., Dutcher, J., Kusmierski, S., Creswell, J.D., Akpan, E., Hallion, L.S., 2023. Neural correlates of mindful disengagement from worry. Journal of Psychopathology and Clinical Science 132, 38–50. 10.1037/abn0000804

Wu, M., Mennin, D.S., Ly, M., 2019. When worry may be good for you: Worry severity and limbic-prefrontal functional connectivity in late-life generalized anxiety disorder. J Affect Disord. Oct 257, 650–657. 10.1016/j.jad.2019.07.022

